# Causal roles of phenotypic age acceleration and metabolic health on dementia: a Mendelian randomisation and structure learning study

**DOI:** 10.64898/2026.09.01.26360731

**Authors:** Alexandra Baousi, Katrina Dobinda, Jingqi Zhu, Xinzhu Yu, Kenneth R. Muir, Artitaya Lophatananon, Brian McMillan, Paul Clarkson, Eugene Y. H. Tang, Hui Guo

## Abstract

**Background:** Phenotypic age acceleration (PhenoAgeAccel), derived from PhenoAge, and MetaboHealth are composite exposures of biological ageing and metabolic health associated with dementia-related outcomes. Whether these associations are causal and reflect the exposures, constituent biomarkers, or both remains unclear.

**Methods:** This study included UK Biobank participants of White British genetic ancestry. MetaboHealth was derived from nuclear magnetic resonance (NMR) metabolomics and PhenoAgeAccel from clinical biomarkers and chronological age. Genome-wide association studies (GWAS) were conducted for MetaboHealth (n=272 568) and PhenoAgeAccel (n=274 077). Independent genome-wide significant variants were used as genetic instruments in two-sample Mendelian randomisation (MR) with FinnGen all-cause dementia summary statistics. Inverse-variance weighting was the primary MR method. Causal network analysis estimated relationships among constituent biomarkers and dementia.

**Findings:** GWAS identified 126 and 141 independent genome-wide significant variants for MetaboHealth and PhenoAgeAccel, of which 109 and 141 were retained as genetic instruments. MR found no evidence of a causal effect of genetically predicted MetaboHealth (per unit: OR 0·83, 95% CI 0·49–1·42; p=0·51) or PhenoAgeAccel (per year: OR 0·99, 95% CI 0·95–1·02; p=0·44) on all-cause dementia, with consistent findings across sensitivity analyses and robust MR methods. Lower lymphocyte percentage and higher NMR-derived glucose had direct relationships with dementia in the joint constituent-biomarker network.

**Interpretation:** MR provided no evidence that either composite exposure causally influenced dementia. The network prioritised lymphocyte percentage and NMR-derived glucose, supporting examination of composite exposures alongside their constituent biomarkers.

**Funding:** NIHR, UKRI, MRC, UK Dementia Research Institute, Innovate UK, and European Union. Full funding details are provided in the acknowledgements.

**Research in context:** *Evidence before this study:* We searched PubMed from database inception to July 11, 2026, without language restrictions, using combinations of the terms “dementia”, “Alzheimer’s disease”, “vascular dementia”, “biological ageing”, “phenotypic age”, “PhenoAge”, “PhenoAgeAccel”, “MetaboHealth”, “metabolomics”, “genome-wide association study”, “Mendelian randomisation”, and “causal network”. Previous GWAS characterised the genetic architecture of PhenoAgeAccel and MetaboHealth. Longitudinal studies linked PhenoAgeAccel, derived from PhenoAge, to incident all-cause, young-onset, and late-onset dementia, dementia subtypes, cognition, and brain structure; MetaboHealth was associated with poorer cognitive performance, 10-year cognitive decline, and reduced functional independence. Related PhenoAge-based measures have also been studied in relation to modifiable factors and intervention response. Genetic causal evidence remained limited: one two-sample MR study found no evidence that PhenoAgeAccel affected Alzheimer’s disease or vascular dementia, while previous causal-discovery research placed Phenotypic Age within a dementia network without examining its constituent biomarkers.

*Added value of this study:* To our knowledge, this is the first study to genetically evaluate PhenoAgeAccel and MetaboHealth as composite exposures and then deconstruct their constituent biomarker relationships using causal network analysis in dementia. It also provides the first large-scale GWAS of MetaboHealth using the complete Phase 3 UK Biobank Nightingale dataset. The expanded GWAS identified additional loci and enlarged the available genetic instrument sets, while the network analysis resolved the constituent biomarker structure underlying both composite exposures.

*Implications of all available evidence:* For causal investigation, analysing PhenoAgeAccel and MetaboHealth alongside their constituent biomarkers reveals relationships obscured within the composite exposures. This does not preclude their use for prediction or risk stratification. Lymphocyte percentage and NMR-derived glucose therefore warrant further investigation as potential indicators of immune and metabolic pathways relevant to dementia.

## Introduction

Dementia comprises neurodegenerative and cerebrovascular disorders that converge clinically in progressive impairment of cognition, behaviour, and everyday function [1]. Because chronological age is non-modifiable and only crudely reflects physiological decline, dementia research has increasingly explored composite measures of biological ageing and metabolic health [2–4]. Among these, MetaboHealth was developed to predict 5- and 10-year all-cause mortality, with higher values indicating a more adverse systemic metabolic profile [5]. Phenotypic Age (PhenoAge) estimates mortality-related biological ageing from chronological age and clinical biomarkers, whereas phenotypic age acceleration (PhenoAgeAccel) denotes deviation from the value expected for chronological age [6]. Unlike CpG-based epigenetic clocks such as DNAm PhenoAge and Horvath’s clock [7], PhenoAgeAccel and MetaboHealth resolve into measured biomarkers amenable to component-level analysis.

In observational studies, higher PhenoAgeAccel was prospectively associated with incident dementia using multivariable Cox regression [8]. Separately, higher MetaboHealth was associated with poorer cognitive performance and faster cognitive decline using multivariable regression and linear mixed-effects models [9,10]. Although these analyses adjusted for measured confounders, residual confounding and reverse causation cannot be excluded, particularly given the prolonged prodromal phase of dementia, supporting the use of complementary genetic analyses [11].

Genome-wide association studies (GWAS) can characterise the genetic architecture of these exposures and provide instruments for Mendelian randomisation (MR) [12,13]. MR uses genetic variants to estimate causal effects with greater robustness to residual confounding and reverse causation than conventional observational analyses [14]. However, when applied to composite exposures, MR estimates their overall effect and cannot distinguish the contributions of individual constituent biomarkers. Null estimates might therefore reflect no overall causal effect, limited statistical power, or heterogeneous constituent relationships that differ in magnitude or direction [15–17].

Network-based methods complement MR by mapping directed relationships among interdependent variables and translating complex clinical constructs into interpretable structures. In previous applications, hyperuricaemia and body-mass index were most proximal to metabolic-syndrome progression [18], whereas dementia-focused causal discovery distinguished variables directly connected to dementia from those operating through intermediate pathways [19].

Yet, to our knowledge, no studies have genetically evaluated PhenoAgeAccel and MetaboHealth as composite exposures before deconstructing their constituent biomarker relationships through causal network analysis. We address this gap by combining GWAS and MR with causal network modelling to evaluate the overall effects of both exposures and map the biological components and pathways underlying their relationships with dementia.

## Methods

This study used individual-level UK Biobank (UKB) data for phenotype derivation, GWAS, and causal network analysis, and FinnGen summary statistics for the all-cause dementia outcome in the two-sample MR analyses. The workflow is shown in figure 1. Figure 1: Analytical framework. UK Biobank participant selection and workflow for exposure derivation, genome-wide association studies, two-sample Mendelian randomisation, and causal network analysis.

**Figure 1:**
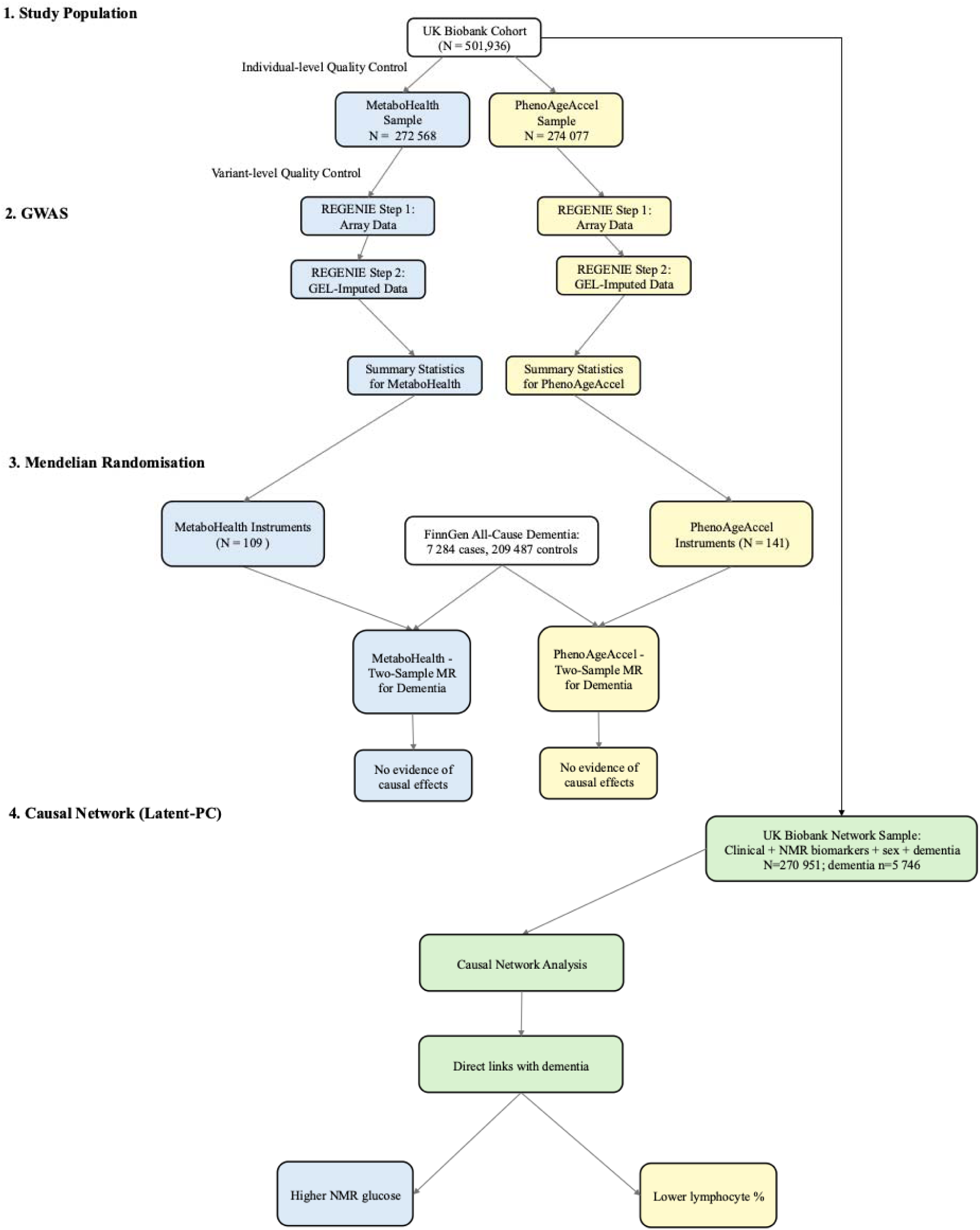
Analytical framework. UK Biobank participant selection and workflow for exposure derivation, genome-wide association studies, two-sample Mendelian randomisation, and causal network analysis.

### Study Design and Participants

UKB is a prospective biomedical cohort study comprising half a million participants recruited between 2006 and 2010, with baseline phenotypic and genotypic data and longitudinal follow-up for health outcomes [20].

Two established exposures were derived using published algorithms: MetaboHealth [5], a composite metabolic health exposure derived from the complete Phase 3 2025 UKB Nightingale nuclear magnetic resonance (NMR) metabolomics dataset, and phenotypic age acceleration (PhenoAgeAccel) [6], calculated as the residual of PhenoAge regressed on chronological age. Exposure derivation is described in detail in the appendix (pp 3–4), including the coefficients used for MetaboHealth and PhenoAgeAccel calculations (appendix pp 16–17).

Individual-level quality control excluded participants for missing genetic principal components, excess genotype missingness or atypical heterozygosity, relatedness, sex-chromosome aneuploidy, or discordance between recorded sex (UKB field 31) and genetically inferred sex (UKB field 22001). In all subsequent UKB analyses, sex denotes recorded sex, which was concordant with genetically inferred sex in all retained participants.

Analyses were further restricted to participants classified within the UKB White British genetic ancestry group to minimise population heterogeneity. Exclusion counts are provided in the appendix (p 7). GWAS models were adjusted for age, sex, genotyping array, and the first ten genetic principal components.

The MetaboHealth GWAS included 272 568 participants with NMR metabolomics data, of whom 99·6% had complete data for all constituent biomarkers. For PhenoAgeAccel derivation, 274 116 participants had chronological age and at least partial data for the required clinical biomarkers; 85·3% had complete data for all biomarkers. After excluding 39 participants with non-finite PhenoAge values, the final sample comprised 274 077 participants. Missingness ranged from 0·001% to 0·196% across MetaboHealth biomarkers and from 2·32% to 12·18% across PhenoAge biomarkers. Missing biomarker values were imputed using chained equations conditional on the remaining biomarkers and covariates [21]. Following suspension of UKB platform access, the first completed dataset containing observed and imputed values was used for downstream analyses.

For two-sample MR, outcome summary statistics were obtained from the European-ancestry FinnGen all-cause dementia GWAS (F5_DEMENTIA; 7 284 cases and 209 487 controls), accessed through OpenGWAS (ID: finn-b-F5_DEMENTIA) [22,23]. Association testing used SAIGE with adjustment for age, sex, genotyping batch, and the first ten genetic principal components. There was no participant overlap between the UKB exposure and FinnGen outcome samples.

### Statistics

#### Genome-wide association study

GWAS of MetaboHealth and PhenoAgeAccel were conducted separately as quantitative traits on the UKB Research Analysis Platform using autosomal variants and two-step REGENIE v2 [24]. Variant quality control used PLINK 2·0 [25]. Step 1 array variants [26] required minor allele frequency (MAF)>0·01, missingness<0·05, and Hardy–Weinberg equilibrium p>1×10 ; step 2 Genomics England-imputed variants [27] required MAF>0·005, missingness<0·05, Hardy–Weinberg equilibrium p>1×10 ¹ , and INFO≥0·8. Indels were removed post-GWAS. Additive models adjusted for age, sex, genotyping array, and ten genetic principal components (appendix p 15); genome-wide significance was p<5×10 . Independent significant variants were identified by linkage disequilibrium (LD) clumping at p<5×10 , r²<0·01, and a 10 Mb window using study-specific LD. Single-nucleotide polymorphism (SNP) heritability and genomic inflation were estimated using LD Score Regression [28] (appendix p 5).

#### Mendelian randomisation analysis

MR reporting followed the STROBE-MR guidance [29]. Two-sample MR analyses were conducted separately for genetically predicted MetaboHealth and PhenoAgeAccel as exposures, with all-cause dementia assessed as the outcome in each analysis using R version 4·6·0. Genome-wide significant, LD-independent variants from the MetaboHealth and PhenoAgeAccel GWAS were used as genetic instruments. Variants with missing reference SNP cluster identifiers (rsIDs) were matched where possible using Ensembl Variant Effect Predictor [30], and unmatched variants were excluded from the MR analysis. Instruments were restricted to biallelic SNPs available in the FinnGen outcome dataset and were matched by exact rsID; proxy variants were not used.

Instrument validity was considered against the three core MR assumptions: relevance, independence from exposure–outcome confounders, and absence of an effect on dementia through pathways other than the exposure [31]. Relevance was assessed using F-statistics. The latter two assumptions cannot be verified directly; potential violations were addressed through population-structure adjustment and investigated using pleiotropy and robust sensitivity analyses.

Exposure and outcome summary statistics were harmonised using the TwoSampleMR 0·7·6 [32], aligning effect alleles and removing non-inferable palindromic variants. Inverse-variance weighting was primary; weighted-median, MR-Egger, simple-mode, and weighted-mode estimates assessed consistency. Cochran’s Q and the MR-Egger intercept assessed heterogeneity and directional pleiotropy. Steiger testing assessed directionality at instrument-set level; leave-one-out and single-variant analyses assessed instrument influence. Robustness to invalid or pleiotropic instruments was evaluated using MR-RAPS[33] and MR-LASSO [34], and the MR-PRESSO global, outlier, and distortion tests. Instrument exclusion occurred only within sensitivity analyses, not the primary analyses. Two-sided p<0·05 was used; power calculations are described in the appendix (p 6).

#### Causal network analysis

Causal network analysis examined the constituent biomarkers of MetaboHealth and PhenoAgeAccel together with chronological age (UKB field 21003), sex (UKB field 31), and all-cause dementia status (field 42018). Dementia status included prevalent and incident records, irrespective of their timing relative to biomarker assessment. The analysis included 270 951 participants with overlapping data across the included variables.

Latent-PC was used to estimate the network structure [35]. The Peter–Clark algorithm [36] identifies relationships that remain after accounting for the other variables and then applies logical rules to infer their likely direction. Latent-PC extends this approach to allow continuous biomarkers and binary variables to be analysed together within a common framework. The analysis assumed that no unmeasured factor jointly influenced the included variables. Continuous variables were standardised before analysis to mirror their scaling within the exposure calculations and account for differences in measurement units. Variable distributions are summarised in the appendix (p 18) as mean (SD) or median (IQR), as appropriate.

A latent correlation matrix was estimated and supplied to the Latent-PC algorithm implemented in the pcalg R package [37]. Fisher’s *Z*-tests were implemented to assess conditional independence relationships, with a prespecified significance threshold of α = 1×10 to favour a sparse network and reduce retention of weakly supported relationships [38]. Background clinical knowledge constrained age and sex as upstream variables and dementia status as a downstream variable, preventing arrows into age or sex or out of dementia without requiring any relationship to be present.

The final network was further annotated using correlation statistics. For each connected pair of variables, Spearman partial correlations were estimated using a conditioning set that included variables neighbouring either member of the pair, accounting for the local network structure surrounding their relationship [39]. Their signs indicated whether associations were positive or negative, while corresponding two-sided p-values tested the null hypothesis of no conditional association. Where no valid conditioning variables were available, marginal Spearman correlations and corresponding p-values were used. Marginal correlations were also calculated for all pairs of variables to describe overall association strength and are presented in the appendix (p 14).

#### Ethics statement

UKB received ethical approval from the North West–Haydock Research Ethics Committee (21/NW/0157); participants provided written informed consent, and data were accessed under application 532708. GWAS used UKB array and Genomics England-imputed genotypes. MR used publicly available FinnGen F5_DEMENTIA summary statistics accessed through OpenGWAS; no FinnGen individual-level data were used.

## Results

### Genome-wide association study

The MetaboHealth GWAS tested 7 253 254 variants after quality control, of which 9 102 reached genome-wide significance (figure 2A). Independent significant variants were annotated to genes using mappings reported in the GWAS Catalog [40]. The strongest signal, rs77303550 on chromosome 16 (p=7·33×10 ²³²), mapped to the *DHODH/HP* region, with neighbouring lead variants mapping to *HPR* and *PKD1L3*. These loci implicated mitochondrial pyrimidine metabolism, haemoglobin scavenging, and acute-phase inflammatory biology [41,42]. On chromosome 15, rs261334 and rs7177289 mapped to the *LIPC/ALDH1A2* region, implicating hepatic lipid and lipoprotein metabolism [43]. The missense variant rs28929474 mapped to *SERPINA1*, linking MetaboHealth variation to protease regulation and hepatic acute-phase biology [44].

**Figure 2:**
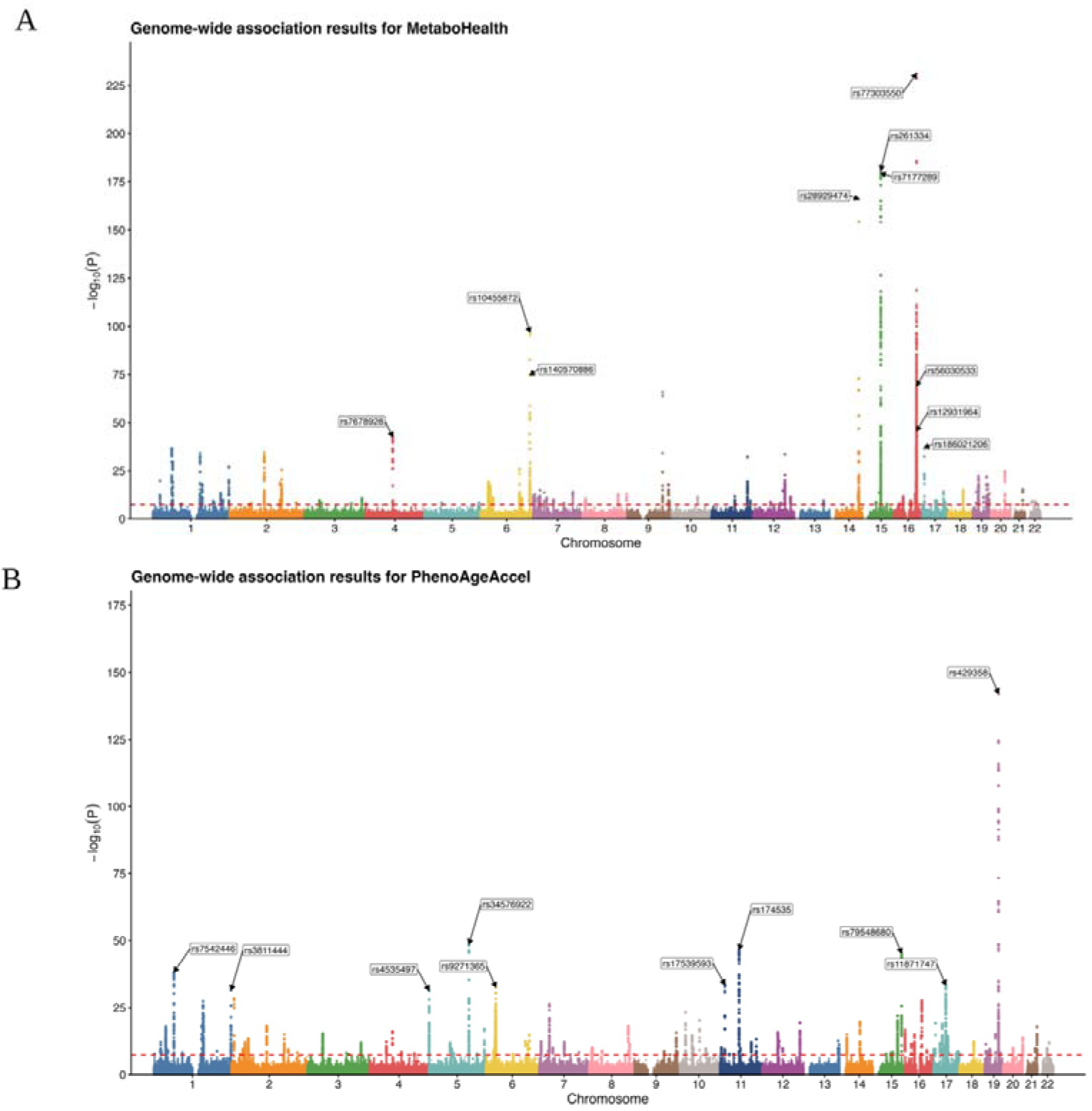
Genome-wide association analyses of MetaboHealth and PhenoAgeAccel. Manhattan plots for MetaboHealth (A; n=272 568) and PhenoAgeAccel (B; n=274 077). The horizontal dashed line denotes the genome-wide significance threshold of p=5×10 . Labels identify independent lead variants after linkage disequilibrium clumping. PhenoAgeAccel=phenotypic age acceleration.

Additional prominent signals at *LPA*, *PPM1K-DT*, and *ASGR1/RPL7AP64* implicated lipoprotein(a)-related transport, branched-chain amino-acid catabolism, and hepatic glycoprotein clearance, respectively [45–47].

The PhenoAgeAccel GWAS tested 7 253 261 variants, of which 10 779 reached genome-wide significance (figure 2B). The strongest signal, rs429358 on chromosome 19 (p=7·53×10 ¹ ³), mapped to *APOE*, the ε4- defining locus central to lipid transport, longevity, and Alzheimer’s disease genetic risk [48]. Further signals included rs34576922 at *SLC12A2-DT/SLC12A2,* rs174535 at *TMEM258/MYRF*, and rs79548680 at *RCCD1*, implicating ion transport, polyunsaturated fatty-acid metabolism, and mitochondrial homeostasis, respectively [49–52]. Additional lead variants included rs3811444 at *TRIM58*, rs4535497 at *SLC12A7*, rs9271365 at *HLA- DQA1/HLA-DRB1*, rs17539593 near *SOX6*, and rs11871747 at *MED24*, further implicating haematological and immune regulation.

### Mendelian randomisation analysis

Two-sample MR included 109 independent genetic instruments for MetaboHealth, all exceeding the conventional F-statistic threshold of 10 (range 30·05–1057·00) [53]. The primary inverse-variance weighted analysis provided no evidence of a causal effect on all-cause dementia (OR 0·83, 95% CI 0·49–1·42; p=0·51; figure 3A-B). Weighted median, MR-Egger, simple-mode, and weighted-mode estimates were also compatible with no effect.

**Figure 3:**
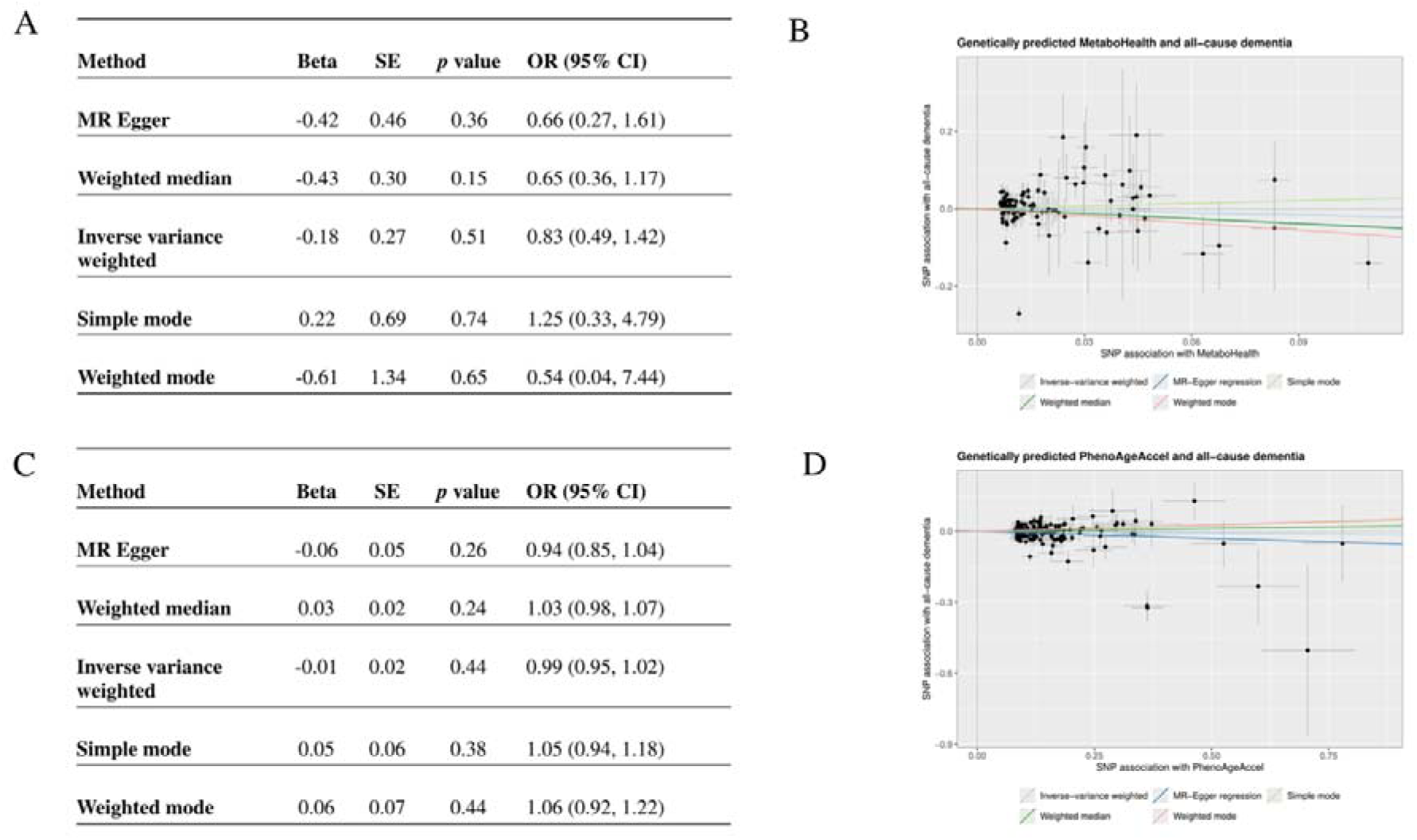
Mendelian randomisation estimates of MetaboHealth and PhenoAgeAccel on all-cause dementia. Tables present method-specific estimates for MetaboHealth (A) and PhenoAgeAccel (C), with corresponding scatter plots showing SNP-specific associations and fitted estimates from each Mendelian randomisation method (B and D, respectively). Horizontal and vertical error bars in the scatter plots represent one SE for the SNP– exposure and SNP–outcome associations. β estimates are presented on the log-odds scale per unit increase in MetaboHealth or per 1-year increase in PhenoAgeAccel; corresponding ORs and 95% CIs are shown in the tables. CI=confidence interval. MR=Mendelian randomisation. OR=odds ratio. SE=standard error. SNP=single- nucleotide polymorphism.

Substantial between-instrument heterogeneity was detected (inverse-variance-weighted Q=288·10; p=2·55×10 ¹ ), although the MR-Egger intercept provided no evidence of directional horizontal pleiotropy (p=0·52). MR-RAPS was also compatible with no effect (OR 1·12, 95% CI 0·75–1·67; p=0·58). Set-level Steiger testing supported the hypothesised direction from MetaboHealth to all-cause dementia (R²=4·01% for the exposure and 0·82% for the outcome; p<0·001). The MR-PRESSO global test indicated outlier-related heterogeneity (p<0·001), and MR-PRESSO and MR-LASSO both identified rs584007 and rs11878597; MR-LASSO retained 107 of 109 instruments for its sensitivity estimate. The resulting sensitivity estimates remained null (MR-PRESSO β=0·064, SE=0·170; p=0·71; MR-LASSO OR 1·07, 95% CI 0·76–1·49; p=0·71), and the MR-PRESSO distortion test was non-significant (p=0·11). Steiger testing was diagnostic, and the MR-PRESSO and MR-LASSO exclusions were confined to sensitivity analyses; no instruments were removed from the primary 109-instrument analysis. Leave-one-out analyses supported the same conclusion, and single-variant forest plots and funnel plots are provided in the appendix (pp 8–10).

The PhenoAgeAccel analysis included 141 independent instruments matched in the FinnGen all-cause dementia outcome data, all exceeding the F-statistic threshold of 10 (range 29·85–218·01). The inverse-variance weighted analysis provided no evidence of a causal effect on all-cause dementia (OR 0·99, 95% CI 0·95–1·02; p=0·44; figure 3C-D). Weighted median, MR-Egger, simple-mode, and weighted-mode estimates were similarly compatible with no effect.

Between-instrument heterogeneity was detected (inverse-variance-weighted Q=226·36; p=5·0×10 ), but the MR-Egger intercept provided no evidence of directional horizontal pleiotropy (intercept=0·0064; SE=0·0071; p=0·37). MR-RAPS was also compatible with no effect (OR 1·00, 95% CI 0·96–1·03; p=0·86). Set-level Steiger testing supported the hypothesised direction from PhenoAgeAccel to all-cause dementia (R²=2·94% for the exposure and 0·64% for the outcome; p=2·78×10 ²² ). The MR-PRESSO global test indicated outlier-related heterogeneity (p<0·001) and identified three outlying variants, while MR-LASSO excluded eight potentially invalid instruments and retained 133 of 141. Following correction, both estimates remained null (MR-PRESSO β=0·004, SE=0·015; p=0·82; MR-LASSO OR 1·00, 95% CI 0·98–1·03; p=0·76), and the MR-PRESSO distortion test was non-significant (p=0·10). Leave-one-out analyses indicated that the overall result was not driven by any single instrument, and single-variant forest plots and funnel plots are provided in the appendix (pp 11–13).

Post-hoc calculations indicated 80% power to detect ORs of at least 1·17 or at most 0·83 for MetaboHealth, and at least 1·20 or at most 0·81 for PhenoAgeAccel (appendix p 6). The analyses therefore had sufficient power to detect moderate effects, although smaller effects could not be excluded.

### Causal network analysis

In the Latent-PC network, age and sex had directed relationships with all-cause dementia, with their direction towards dementia imposed a priori based on established epidemiological evidence [54,55]. The network also identified directed relationships from lymphocyte percentage and NMR-derived glucose to dementia (figure 4). The presence of these relationships was determined from the data, whereas their orientation towards dementia followed the prespecified downstream constraint. The directed relationship from lymphocyte percentage to dementia had a negative partial correlation (−0·0180; p=5·38×10 ²¹), whereas the directed relationship from NMR-derived glucose to dementia had a positive partial correlation (0·0127; p=3·52×10 ¹¹). Thus, the dementia-facing biomarker neighbourhood was characterised by lower relative lymphocyte percentage and higher NMR-derived glucose after conditioning on the surrounding network structure.

**Figure 4:**
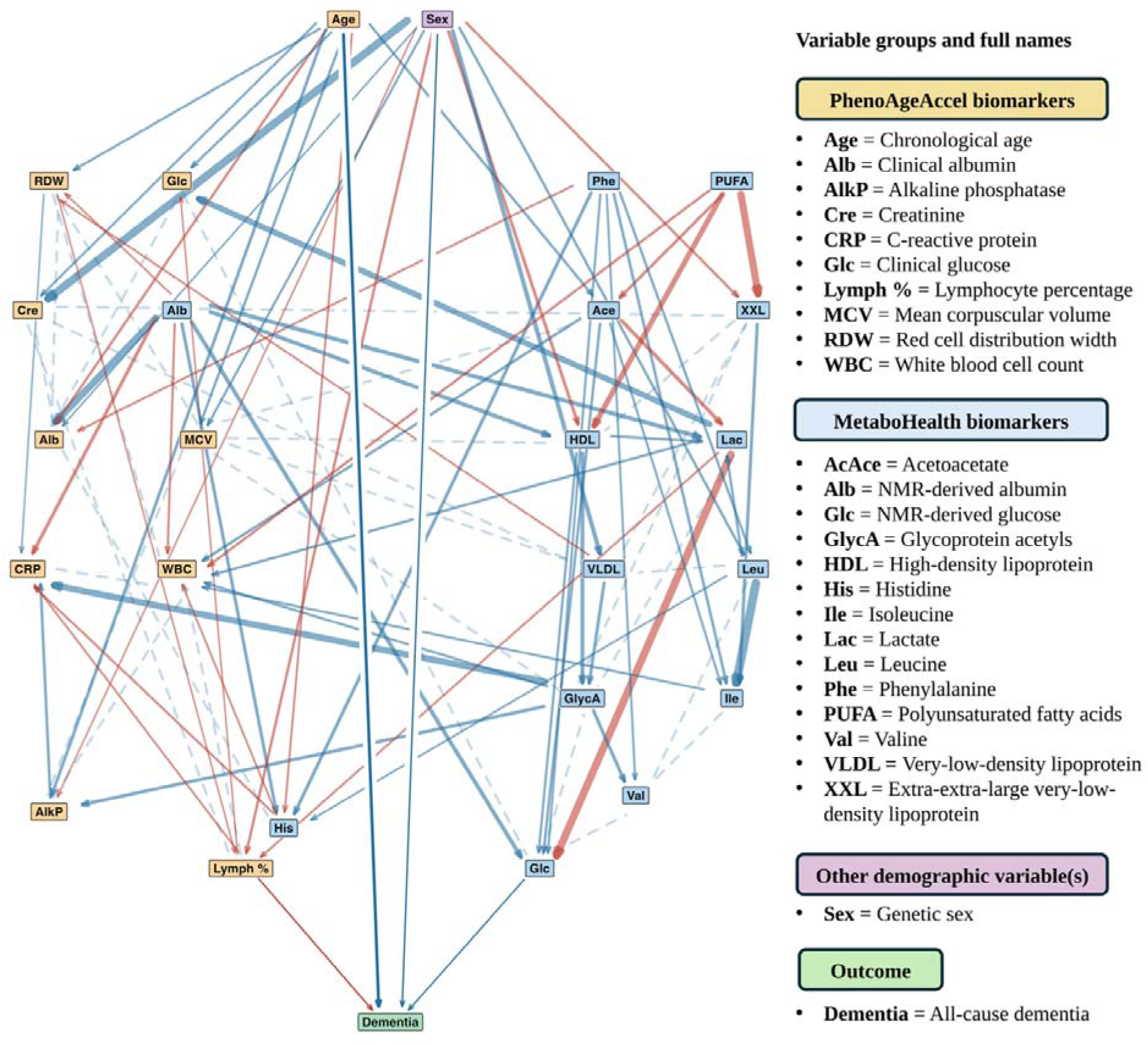
Causal network of individual PhenoAgeAccel and MetaboHealth biomarkers with all-cause dementia. The inferred network shows relationships among PhenoAgeAccel and MetaboHealth constituent biomarkers and all-cause dementia. Age and sex were constrained upstream and dementia downstream; these constraints affected the orientation of relationships but did not require any relationship to be present. Arrows denote directed relationships, while dashed lines denote relationships for which direction could not be determined. Blue and red denote positive and negative partial correlations, respectively, and arrow thickness is proportional to the magnitude of the partial correlation. Full biomarker names are provided in the figure key.

The network also recovered a broader ancestral structure linking lactate with both glucose measures, glycoprotein acetyls with C-reactive protein, polyunsaturated fatty acids with very-low-density lipoprotein (VLDL) and high-density lipoprotein (HDL) measures, leucine with isoleucine, and sex with creatinine. All displayed variables formed part of the ancestral network of dementia, through either direct relationships or pathways involving intermediate variables.

This topological specificity contrasted sharply with the broader, highly heterogeneous correlation structure of the underlying constituent biomarkers. MetaboHealth contained strongly correlated biomarker clusters, particularly average VLDL particle diameter with extra-extra-large VLDL particle lipids (r=0·91), and branched-chain amino acids, including isoleucine–leucine (r=0·89), leucine–valine (r=0·88), and isoleucine–valine (r=0·82; appendix p 14). Strong inverse correlations were also observed between polyunsaturated fatty acids and extra-extra-large VLDL particle lipids (r=−0·79), average VLDL particle diameter (r=−0·73), and glycoprotein acetyls (r=−0·50). Matched clinical and NMR-derived measures were moderately correlated for glucose (r=0·56) and albumin (r=0·51).

## Discussion

The principal finding was a divergence between composite- and biomarker-level analyses. Genetically predicted MetaboHealth and PhenoAgeAccel were not associated with all-cause dementia, whereas, among constituent biomarkers, only lymphocyte percentage and NMR-derived glucose were directly connected to dementia. Neither exposure therefore showed evidence of a unitary causal effect.

The lack of evidence for a composite causal effect does not detract from the value of MetaboHealth and PhenoAgeAccel as measures of systemic health. By summarising information across physiological systems, composite scores reduce dimensionality and capture multisystem vulnerability beyond individual biomarkers. MetaboHealth was associated with mortality across 12 cohorts and improved long-term mortality prediction relative to conventional risk factors in an external validation cohort, while Phenotypic Age differentiated morbidity and mortality risk beyond chronological age across diverse population groups [5,56]. Subsequent studies have supported their prognostic value, although improvements over established models have generally been modest [57–59].

However, aggregation limits causal interpretation. Similar scores can arise from different biomarker profiles, and weights optimised for mortality prediction do not imply a shared pathway or direction of effect on dementia. Genetic variants may influence a composite through different constituents, causing heterogeneous or opposing effects to be averaged in MR. Clinical transferability also depends on validation across populations, measurement platforms, and repeated assessments [60]. The present findings therefore distinguish the established predictive utility of these scores from their more limited interpretation as mechanistically coherent or directly modifiable exposures.

Previous GWAS have characterised the genetic architecture of MetaboHealth and PhenoAgeAccel [13,61–63] and our larger analyses largely reinforced these findings. For PhenoAgeAccel, rs429358 at *APOE* and rs3811444 at *TRIM58* replicated previously reported lead associations, while the *FADS1/FADS2* and *SLC12A7* regions were represented by neighbouring variants [13]. Additional associations in the present study at *SLC12A2, RCCD1, HLA-DQA1/HLA-DRB1*, *SOX6*, and *MED24* broadened the candidate architecture. For MetaboHealth, identical lead associations at *SERPINA1* and *LPA*, together with recurrence of the *DHODH–HP– HPR, LIPC–ALDH1A2*, and *PPM1K* regions, corroborated previous evidence [12] implicating hepatic lipid metabolism, acute-phase biology, and branched-chain amino-acid metabolism. Overall, the present GWAS extended rather than contradicted earlier work, providing greater genomic resolution and larger instrument sets for subsequent causal analysis.

Within the network, lower lymphocyte percentage was related to dementia. This is consistent with prospective evidence associating lower lymphocyte counts and an imbalance between innate and adaptive immunity with increased dementia risk, although cross-sectional data from adults aged 90 years and older associated higher absolute lymphocyte counts with prevalent dementia [64,65]. These findings are not directly comparable because lymphocyte percentage is a relative measure: it can decrease through fewer lymphocytes, increases in other white-cell populations, or both. The relationship may therefore reflect immune ageing or chronic systemic inflammation rather than a direct effect of lymphocyte abundance.

Higher NMR-derived glucose may similarly represent broader glycaemic dysfunction associated with insulin resistance, diabetes, and adiposity. Diabetes and midlife obesity are established modifiable dementia risk factors [66], although genetic evidence varies by outcome. Genetically higher plasma glucose has been associated with unspecified dementia but not Alzheimer’s disease or vascular dementia [67]; two-sample MR of type 2 diabetes liability was null for Alzheimer’s disease [68], whereas one-sample MR reported modest associations with all-cause, vascular, and Alzheimer’s dementia [69]. Similarly, body-mass index showed no MR association with Alzheimer’s disease but was subsequently associated with vascular-related dementia [70]. The glucose relationship may therefore reflect metabolic and vascular pathways contributing to all-cause dementia. Together, the network findings prioritise immune and glycaemic pathways for further investigation without identifying lymphocyte percentage or circulating glucose as direct intervention targets.

Latent-PC distinguished biomarkers related to dementia from those connected through other measures; for example, routine glucose was linked to dementia through NMR-derived glucose rather than independently [35]. Composite scores were excluded because including them alongside their constituent biomarkers could introduce relationships determined by score construction rather than underlying biology. This could violate the faithfulness assumption, which requires patterns of conditional association and independence in the data to reflect the underlying causal structure [71].

Furthermore, network interpretation depends on the variables and assumptions applied. Latent-PC also assumes causal sufficiency, meaning that the principal shared causes of the analysed variables are represented [36], although unmeasured influences from lifestyle, medication, or comorbidity may remain. This concern was partly addressed by modelling age, sex, and the wider biomarker structure jointly and by applying prespecified temporal constraints. MR provided a complementary analysis less susceptible to conventional confounding and reverse causation [72], but evaluated the composite exposures rather than the individual network relationships. The methods therefore triangulated evidence across different analytical levels rather than replicating the same causal question. The small partial correlations also require cautious clinical interpretation and replication using longitudinal data and alternative structure-learning approaches.

The broader design was strengthened by strong genetic instruments, non-overlapping exposure and outcome samples, and broadly consistent estimates across sensitivity analyses and robust MR methods. Restriction to White British participants reduced bias from population structure, while the use of all-cause dementia increased power to detect relationships shared across dementia presentations. The network was derived from a single cohort, and replication in independent and ancestrally diverse populations, alongside dementia subtype-specific analyses, will be important for establishing generalisability.

Taken together, these findings support a shift from treating composite health scores as single causal exposures towards investigating the biomarker pathways they summarise, providing a more specific basis for research into dementia mechanisms and prevention.

## Contributors

Conceptualisation, AB, JZ, KD, XY, HG, KRM, PC, BM, EYHT, AL; data analysis, AB, JZ, KD; funding acquisition, HG, KRM, PC, BM, EYHT, AL; supervision, KRM, AL, PC, BM, EYHT and HG; visualisation, AB and KD; writing – original draft, AB; and writing – review & editing, AB, KD, JZ, XY, KRM, AL, PC, BM, EYHT and HG. AB, KD and JZ accessed and verified the underlying data. All authors read and approved the final version of the manuscript.

## Data Availability

Summary statistics from the GWAS for PhenoAgeAccel and MetaboHealth will be deposited in the GWAS Catalog and made available upon publication. Individual-level UKB data used in this study are available to researchers via application to UKB (www.ukbiobank.ac.uk), subject to institutional and ethical approvals. Code for all statistical analyses is available on request.

## Acknowledgements

This research has been conducted using the UKB Resource under Application Number 532708. The authors are grateful to the participants and investigators of both the UKB and FinnGen studies, whose invaluable contributions made this research possible. We also thank Sinan Shi (University of Oxford) for guidance regarding the GEL-imputed UKB data, and Xiaoguang Xu and David Scannali (University of Manchester) for advice on the GWAS methodology.

AB, HG, KRM, AL, BM, PC and EYHT acknowledge funding support from the National Institute for Health and Care Research (NIHR) School for Social Care Research, on behalf of the NIHR Three Schools’ Dementia Research Programme, and the NIHR Research Programme for Social Care [NIHR-SSCR-DS07].

HG, KD and BM acknowledge support from the UKRI AI Programme and the Engineering and Physical Sciences Research Council for CHAI (Causality in Healthcare AI Hub) [grant number EP/Y028856/1].

JZ was supported by the Medical Research Council [grant number: MR/W007428/1].

XY received support from the UK Dementia Research Institute at Imperial College London (MC_PC_17114) and NIHR Imperial Biomedical Research Centre (BRC).

KRM, AL and BM acknowledge funding from the European Union’s funded projects COMFORTage (Horizon Europe research and innovation program under grant agreement no. 101137301) and are further supported by Innovate UK under grant agreement no. 10103541.

EYHT was funded by an NIHR Advanced Fellowship [NIHR304435] for this study.

The views and opinions expressed are, however, those of the authors only and do not necessarily reflect those of the NIHR, the Department of Health and Social Care, UKRI, the Medical Research Council, the European Union, or the European Health and Digital Executive Agency (HaDEA). Neither the European Union, HaDEA, nor any other granting authority can be held responsible for them.

