## appendix for "Causal roles of phenotypic age acceleration and metabolic health on dementia: a Mendelian randomisation and structure learning study"

### Supplementary Methods

#### Supplementary Method 1: Scoring Algorithm for MetaboHealth [1]

MetaboHealth was derived from 14 circulating biomarkers measured using the Nightingale nuclear magnetic resonance (NMR) metabolomics platform. The constituent biomarkers were glucose, lactate, histidine, isoleucine, leucine, valine, phenylalanine, acetoacetate, albumin, glycoprotein acetyls, the ratio of polyunsaturated fatty acids to total fatty acids, total lipids in extremely large very-low-density lipoprotein particles, total lipids in small high-density lipoprotein particles, and very-low-density lipoprotein particle diameter.

Biomarker concentrations were log transformed after adding a constant of 1 and were subsequently standardised to a mean of 0 and standard deviation of 1 within the analytical sample. The MetaboHealth score was calculated as:

$$\begin{aligned} \begin{aligned} MetaboHealth\_Score= \sum\left( wᵢ \times z-score\left( log\left( metaboliteᵢ + 1 \right) \right) \right) \#\#\# \end{aligned}\# \end{aligned}$$

where metaboliteᵢ represents the concentration of the ith biomarker, z-score denotes standardisation of the log-transformed biomarker concentration, and wᵢ represents the corresponding published weight for that biomarker. The weighted values were summed across all 14 biomarkers to generate a continuous score for each participant. The published biomarker weights were applied without re-estimation in the present sample. Higher MetaboHealth scores indicated a less favourable metabolic profile and greater mortality-associated metabolic risk. In the analytical sample, MetaboHealth scores were centred at zero and ranged from −1·90 to 4·29.

#### Supplementary Method 2: Scoring Algorithm for PhenoAgeAccel [2]

PhenoAge was derived using chronological age and nine clinical biomarkers: albumin, creatinine, glucose, C-reactive protein, lymphocyte percentage, mean cell volume, red cell distribution width, alkaline phosphatase, and white blood cell count.

PhenoAge translates an individual's mortality risk, derived from a Gompertz proportional hazards model that combines chronological age and nine clinical biomarkers, into biological age in years:

$$\begin{aligned} \begin{aligned} PhenoAge = 141\cdot50225 + \frac{\ln\left( -0\cdot00553 \times\ln\left( 1 - mortality_{risk} \right) \right)}{0\cdot090165}\#\# \end{aligned}\# \end{aligned}$$

Where the mortality risk is computed using the Gompertz model:

$$mortality\_risk = 1 - exp( -exp(xb) \times\frac{(exp\left( 120 \times\gamma\right)-1)}{\gamma}$$

With γ = 0⋅0076927 (the Gompertz hazard slope) and $xb$ representing the linear predictor:

$$\begin{aligned} xb = -19\cdot9067 \left( constant \right)+ \sum_{i=1}^{9} w_{i}x_{i} \#\#\#\# \end{aligned}$$

Where $w_{i}$ are the respective weights and $x_{i}$ are the biomarker values, including chronological age.

To derive phenotypic age acceleration (PhenoAgeAccel), PhenoAge was regressed on chronological age:

$${PhenoAge}_{i}=\alpha+\beta({Age}_{i})+\varepsilon_{i}$$

PhenoAgeAccel was then defined as the residual from this model:

$${PhenoAgeAccel}_{i}=\varepsilon_{i}$$

This residualised measure was used for genome-wide analysis to capture variation in biological ageing that was not explained by chronological age alone. Positive PhenoAgeAccel values indicate older biological age than expected for chronological age, whereas negative values indicate younger biological age than expected. In the analytical sample, mean PhenoAge was 50·81 years, with observed values from 18·14 to 123·83 years. PhenoAgeAccel was centred on zero and spanned -19·51 to 76·22 years.

#### Supplementary Method 3: LD score regression results for MetaboHealth and PhenoAgeAccel GWAS

LD score regression was performed using the Bulik-Sullivan LDSC software package [3]. GWAS summary statistics were analysed against 1000 Genomes Phase 3 LD score reference data and HapMap3 regression weights, excluding the Major Histocompatibility Complex (MHC) region [4]. This was omitted to minimise potential bias arising from its complex linkage disequilibrium structure and dense concentration of variants involved in immune function, which can disproportionately affect genome-wide heritability and inflation estimates [5].

For MetaboHealth, the analysis included 822 779 SNPs from the GWAS summary statistics, of which 812 325 remained after merging with the LD score reference panel and regression weights. The observed-scale SNP heritability was 0⋅0759 (SE = 0⋅0090), indicating a detectable common-variant genetic contribution to MetaboHealth. The genomic control inflation factor was λ_GC_ = 1.3546 and the mean χ² statistic was 1.5899, consistent with inflation of association statistics. The LDSC intercept was 1⋅1186 (SE = 0⋅0136), suggesting modest residual inflation beyond that expected from polygenic signal alone. The attenuation ratio was 0⋅2011 (SE = 0⋅0231), indicating that approximately 20% of the inflation above the null was attributable to confounding or related sources of bias.

For PhenoAgeAccel, 822 780 SNPs were included in the summary statistics, of which 812 326 remained after merging with the LD score reference panel and regression weights. The observed-scale SNP heritability was 0⋅0860 (SE = 0⋅0059), with λ_GC_ = 1⋅4069, mean χ² = 1⋅6513, LDSC intercept = 1⋅1303 (SE = 0⋅0134), and attenuation ratio = 0⋅2001 (SE = 0⋅0206). Similar to MetaboHealth, the results support a measurable common-variant genetic contribution and are consistent with a polygenic architecture [6].

#### Supplementary Method 4: Instrument Strength and Post-Hoc Power Assessment

To evaluate the individual and collective vulnerability of the genetic instruments to weak instrument bias, individual variant F-statistics were computed using the equation $F=(\beta_{exposure}/{SE}_{exposure})$^2^ and the cumulative proportion of phenotypic variance explained by the genetic instrument sets ${(R}_{xz}^{2})$ was calculated as the sum of individual variant variances derived from $R_{snp}^{2}=F/(F+ N_{exposure}-2)$ [7]

For the MetaboHealth exposure, assuming a baseline sample scale of N= 272 568, a final set of 109 independent genetic variants was successfully retained. Individual variant metrics demonstrated robust instrument strength, yielding a minimum single-variant F-statistic of 30⋅05 and a maximum of 1057⋅00, significantly exceeding the conventional weak-instrument threshold (*F* < 10) [8]. Cumulatively, this genetic instrument set explained approximately 4⋅01% of the overall variance in MetaboHealth ${(R}_{xz}^{2}= 0\cdot04005).$

For the PhenoAgeAccel exposure, assuming a baseline sample scale of N= 274 077, a separate set of 141 independent genetic instruments was successfully matched. Individual instrument profiles were similarly robust, yielding a minimum variant F-statistic of 29⋅85 and a maximum of 218⋅01. Cumulatively, the total phenotypic variance explained by the PhenoAgeAccel genetic instrument set was calculated at 2⋅94% ${(R}_{xz}^{2}= 0\cdot02940).$

Post-hoc analytical power and the total sample sizes required to achieve 80% power were calculated using the FinnGen all-cause dementia sample characteristics and a two-sided significance threshold of α=0·05. Power was estimated analytically under a non-central chi-square framework, accounting for the proportion of exposure variance explained by the genetic instruments and a range of prespecified causal odds ratios [9]. The observed standard errors of the MR estimates were not incorporated into these calculations and are reflected separately in the reported confidence intervals.

For MetaboHealth, the genetic instruments explained 4·01% of exposure variance. In the available outcome sample (216 771 participants; 7 284 cases and 209 487 controls; case fraction 3·36%), the minimum detectable ORs at 80% power were 1·168 for a risk-increasing effect and 0·834 for a protective effect. For PhenoAgeAccel, the genetic instruments explained 2·94% of exposure variance. In the available outcome sample, the minimum detectable ORs at 80% power were 1·196 for a risk-increasing effect and 0·807 for a protective effect.

### Supplementary Figures

#### Supplementary Figure 1: Selection of UK Biobank participants for the MetaboHealth and PhenoAgeAccel genome-wide association studies

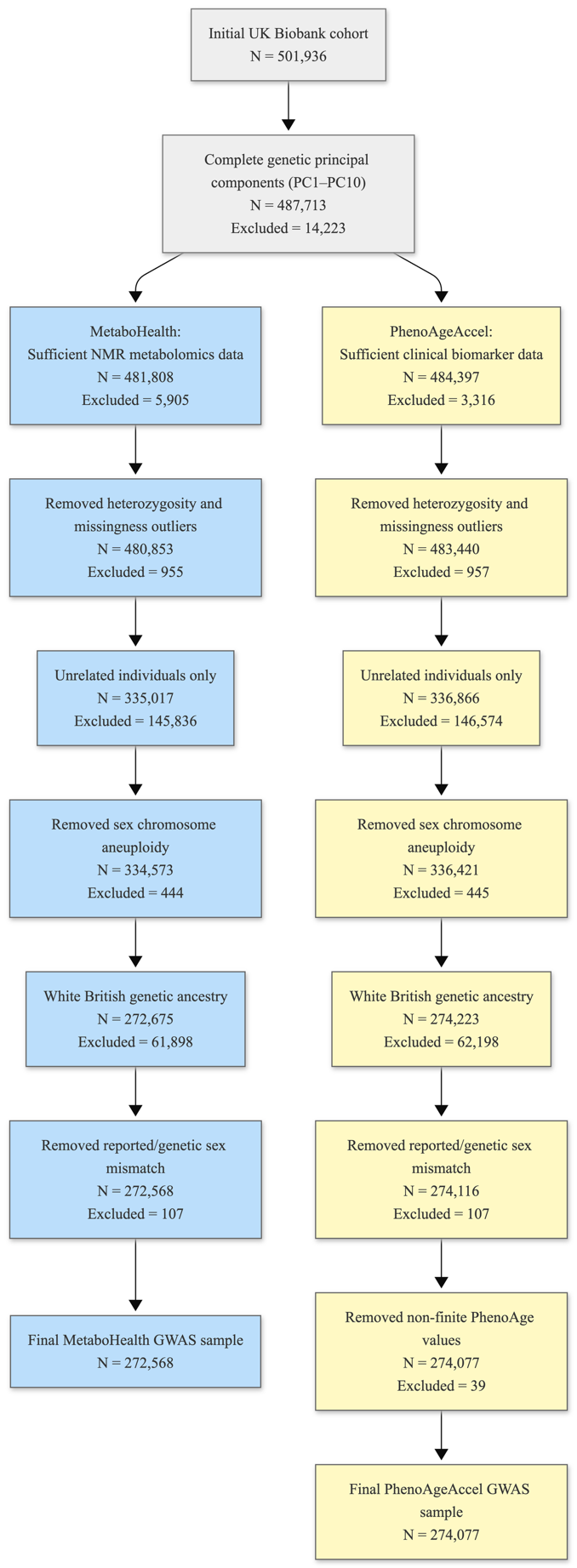

#### Supplementary Figure 2: Funnel plot of single-variant Mendelian randomisation estimates for MetaboHealth and all-cause dementia

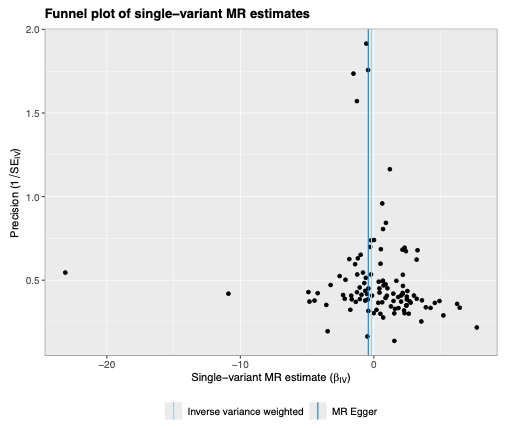

#### Supplementary Figure 3: Forest plot of single-variant Mendelian randomisation estimates for MetaboHealth and all-cause dementia

Single-variant MR estimates are shown with IVW and MR-Egger summary estimates; horizontal lines represent 95% confidence intervals.

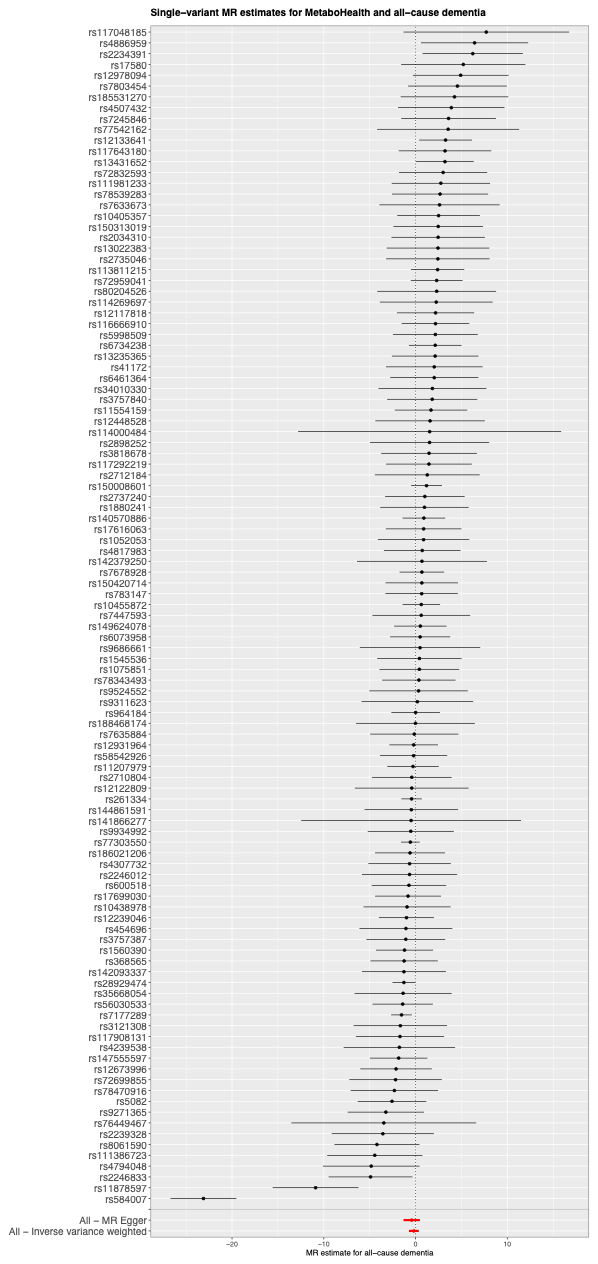

#### Supplementary Figure 4: Leave-one-out sensitivity analysis for MetaboHealth and all-cause dementia

MR estimates are shown after sequential exclusion of each genetic instrument. The “All” row represents the estimate using all instruments.

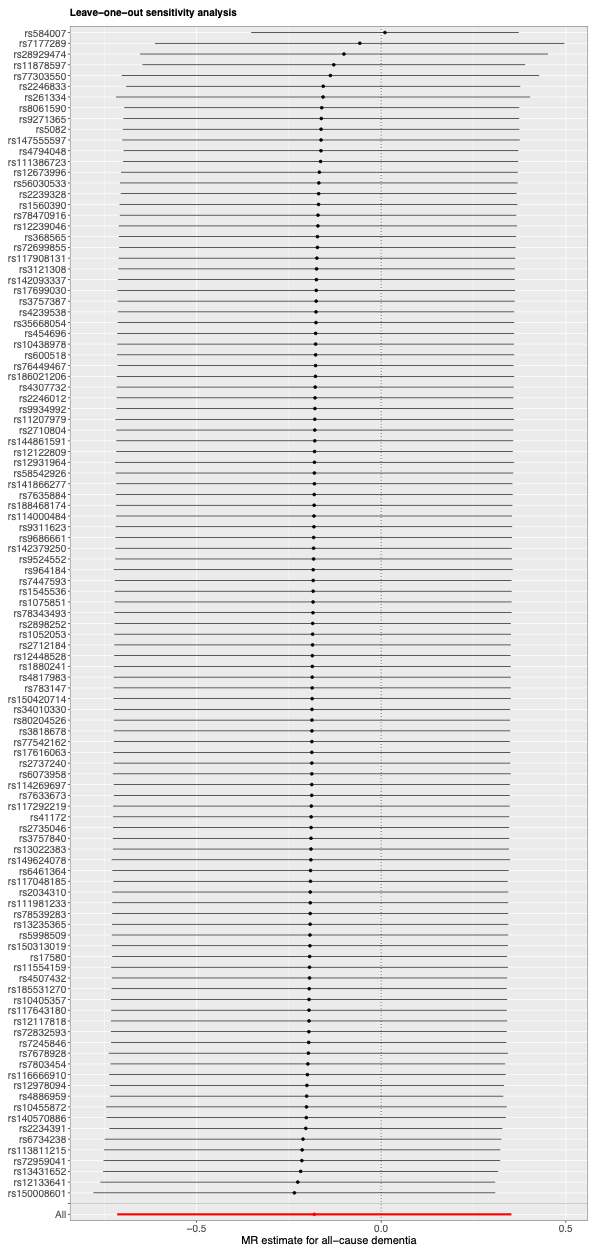

#### Supplementary Figure 5: Funnel plot of single-variant Mendelian randomisation estimates for PhenoAgeAccel and all-cause dementia

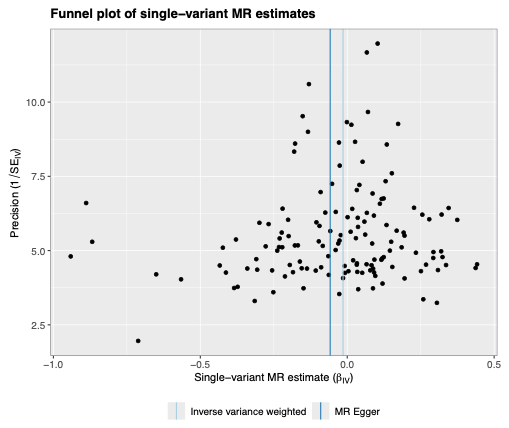

#### Supplementary Figure 6: Forest plot of single-variant Mendelian randomisation estimates for PhenoAgeAccel and all-cause dementia

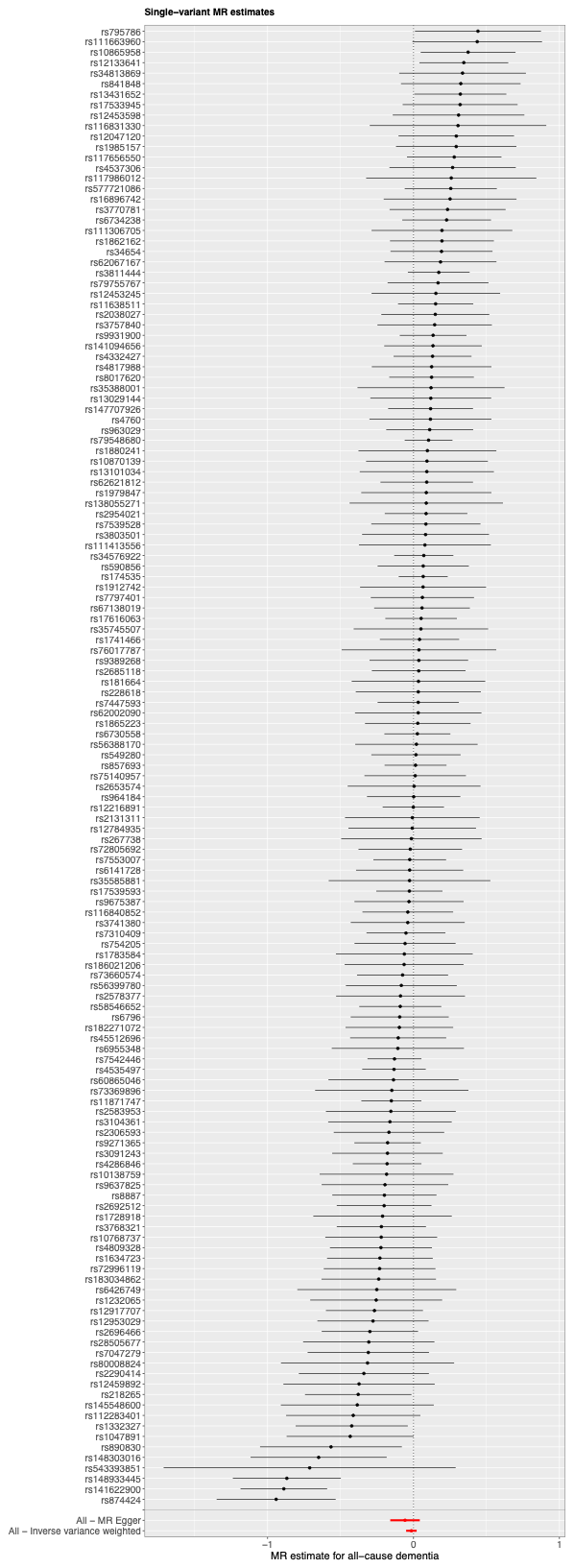

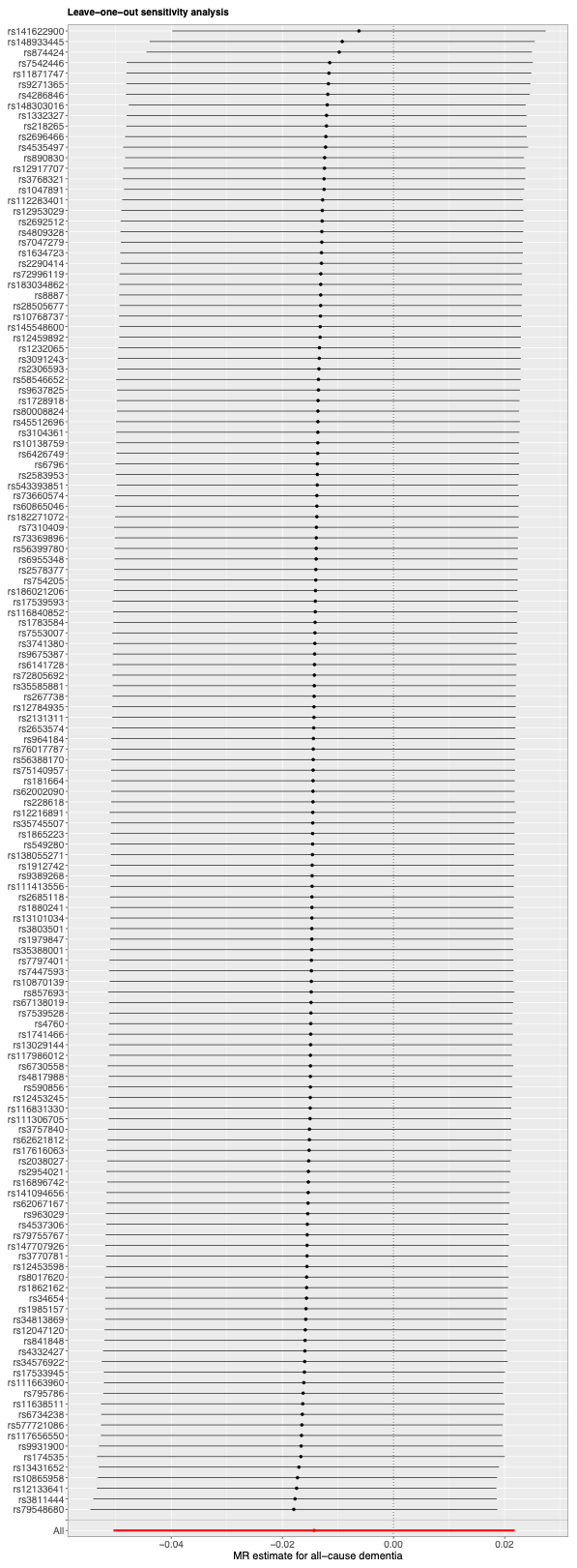

#### Supplementary Figure 7: Leave-one-out sensitivity analysis for PhenoAgeAccel and all-cause dementia

#### Supplementary Figure 8: Pairwise Spearman correlation matrix of MetaboHealth and PhenoAgeAccel constituent biomarkers

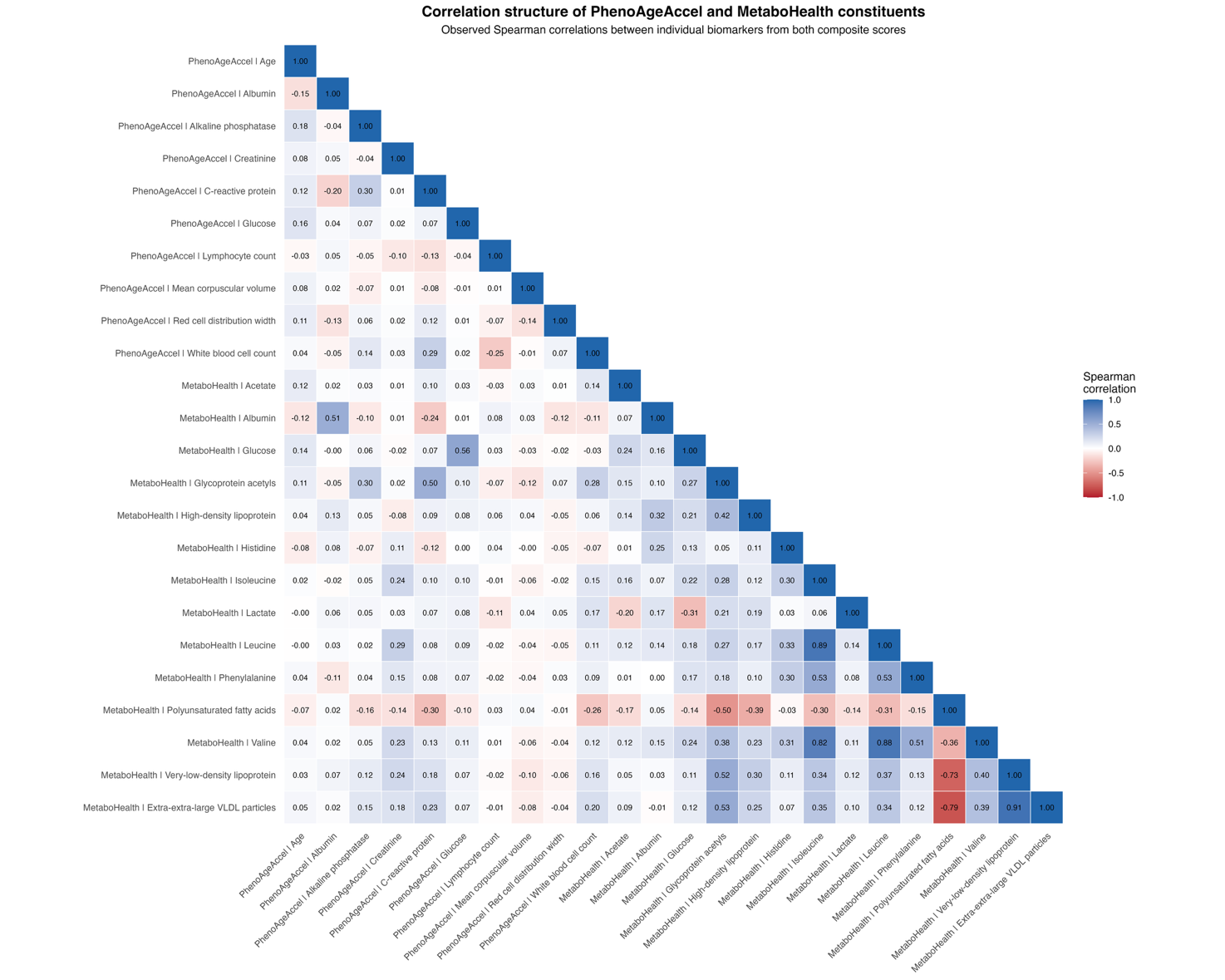

### Supplementary Tables

#### Supplementary Table 1: Definitions of covariates from the UK Biobank

|  | **Variables** | **UKB Showcase field ID** | **Note** |
| --- | --- | --- | --- |
| Covariates | Age | 21003 | Age at recruitment; derived integer years at baseline assessment. |
|  | Recorded Sex | 31 | Sex obtained from the central registry at recruitment and updated where participants reported it to be incorrect; 0 = female, 1 = male. |
|  | Genotype Array | 22000 | Genotype measurement batch; distinguishes UK BiLEVE and UK Biobank Axiom arrays. |
|  | Principal Components 1-10 | 22009 | Genetic principal components; first 10 used to account for population structure |

#### Supplementary Table 2: UK Biobank variables and published coefficients used to derive MetaboHealth

| **Component** | **Abbreviation** | **UKB Showcase field ID** | **Native UKB unit** | **Published MetaboHealth Weight** |
| --- | --- | --- | --- | --- |
| Total lipids in chylomicrons and extremely large very-low-density lipoprotein particles | XXL-VLDL-L | 23482 | mmol/L | −0⋅120 |
| Total lipids in small high-density lipoprotein particles | S-HDL-L | 23573 | mmol/L | 0⋅152 |
| Average diameter of very-low-density lipoprotein particles | VLDL-D | 23431 | nm | 0⋅124 |
| Polyunsaturated fatty acids to total fatty acids ratio | PUFA/FA | 23453 | % | −0⋅149 |
| NMR-derived glucose | Glucose | 23470 | mmol/L | 0⋅099 |
| Lactate | Lactate | 23471 | mmol/L | 0⋅106 |
| Histidine | Histidine | 23463 | mmol/L | −0⋅123 |
| Isoleucine | Isoleucine | 23465 | mmol/L | 0⋅092 |
| Leucine | Leucine | 23466 | mmol/L | 0⋅094 |
| Valine | Valine | 23467 | mmol/L | 0⋅115 |
| Phenylalanine | Phenylalanine | 23468 | mmol/L | 0⋅139 |
| Acetoacetate | Acetoacetate | 23476 | mmol/L | 0⋅070 |
| NMR-derived albumin | Albumin | 23479 | g/L | −0⋅168 |
| Glycoprotein acetyls | GlycA | 23480 | mmol/L | 0⋅141 |

#### Supplementary Table 3: UK Biobank variables and published coefficients used to derive PhenoAgeAccel

| **Component** | **UKB Showcase field ID** | **Native UKB unit** | **Published PhenoAge Weight** |
| --- | --- | --- | --- |
| Chronological age at assessment | 21003 | Years | 0⋅0804 |
| Albumin, clinical biochemistry | 30600 | g/L | −0⋅0336 |
| Creatinine | 30700 | µmol/L | 0⋅0095 |
| Glucose, clinical biochemistry | 30740 | mmol/L | 0⋅1953 |
| C-reactive protein | 30710 | mg/L | 0⋅0954 |
| Lymphocyte percentage | 30180 | % | −0⋅0120 |
| Mean corpuscular volume | 30040 | fL | 0⋅0268 |
| Red blood cell distribution width | 30070 | % | 0⋅3306 |
| Alkaline phosphatase | 30610 | U/L | 0⋅0019 |
| White blood cell count | 30000 | 10⁹ cells/L | 0⋅0554 |

#### Supplementary Table 4: Descriptive characteristics of constituent biomarkers in the causal-network cohort before standardisation

| **Measure** | **N available** | **Descriptive statistic** |
| --- | --- | --- |
| **All-cause dementia** | 270 951 | 5 743 cases (2·12%), 265 208 controls (97·88%) |
| **Recorded Sex** | 270 951 | 144 410 females (53·30%), 126 541 males (46·70%) |
| Chronological age, years | 270 951 | 56·83 (7·98), mean (SD) |
| Albumin, g/L | 270 951 | 45·26 (2·61), mean (SD) |
| Alkaline phosphatase, U/L | 270 951 | 80·20 (67·10 to 95·60), median (IQR) |
| Creatinine, μmol/L | 270 951 | 70·60 (61·70 to 81·00), median (IQR) |
| C-reactive protein, mg/dL | 270 951 | 0·13 (0·07 to 0·27), median (IQR) |
| Routine clinical glucose, mmol/L | 270 951 | 4·93 (4·60 to 5·31), median (IQR) |
| Lymphocyte percentage, % | 270 951 | 28·61 (7·33), mean (SD) |
| Mean corpuscular volume, fL | 270 951 | 91·36 (4·36), mean (SD) |
| Red cell distribution width, % | 270 951 | 13·32 (12·90 to 13·82), median (IQR) |
| White blood cell count, ×10⁹/L | 270 951 | 6·64 (5·64 to 7·83), median (IQR) |
| Total lipids in chylomicrons and extremely large VLDL particles, mmol/L | 270 951 | 0·17 (0·07–0·33), median (IQR) |
| Total lipids in small HDL particles, mmol/L | 270 951 | 1·19 (0·16), mean (SD) |
| Average diameter of VLDL particles, nm | 270 951 | 38·71 (1·26), mean (SD) |
| Polyunsaturated fatty acids to total fatty acids ratio, % | 270 951 | 42·01 (3·76), mean (SD) |
| Glucose, NMR-derived, mmol/L | 270 951 | 3·58 (3·13–4·06), median (IQR) |
| Lactate, mmol/L | 270 951 | 4·05 (1·15), mean (SD) |
| Histidine, mmol/L | 270 951 | 0·07 (0·06–0·07), median (IQR) |
| Isoleucine, mmol/L | 270 951 | 0·05 (0·04–0·06), median (IQR) |
| Leucine, mmol/L | 270 951 | 0·10 (0·09–0·12), median (IQR) |
| Valine, mmol/L | 270 951 | 0·21 (0·04), mean (SD) |
| Phenylalanine, mmol/L | 270 951 | 0·05 (0·04–0·05), median (IQR) |
| Acetoacetate, mmol/L | 270 951 | 0·01 (0·01–0·02), median (IQR) |
| Albumin, NMR-derived, g/L | 270 951 | 39·79 (3·43), mean (SD) |
| Glycoprotein acetyls, mmol/L | 270 951 | 0·82 (0·12), mean (SD) |

Note: Data are presented as n (%) for all-cause dementia, mean (SD) for approximately symmetric continuous variables, and median (IQR) for asymmetric continuous variables. Constituent biomarkers are presented on their original scales before z-standardisation. IQR=interquartile range; NMR=nuclear magnetic resonance; SD=standard deviation; VLDL=very-low-density lipoprotein.
